# The Etiology of Hypothyroidism Is Revealed by Alternative Genetics Association Study Methodologies

**DOI:** 10.1101/2022.10.04.22280703

**Authors:** Amos Stern, Roei Zucker, Michal Linial

**Author notes:** Corresponding author: Michal Linial, Dept. of Biological Chemistry, Institute of Life Sciences, The Hebrew University of Jerusalem, Jerusalem, 91904, Israel.

## Abstract

Hypothyroidism is a common disorder of the endocrine system in which the thyroid gland does not produce enough thyroid hormones. About 12% of the population in the USA will develop substantial thyroid deficiency over their lifetime, mostly as a result of iodine deficiency. The hypothyroidism phenotype also includes individuals that suffer from thyroid development abnormalities (congenital hypothyroidism, CH). Using a large population study, we aimed to identify the functional genes associated with an increase or decreased risk for hypothyroidism (ICD-10, E03). To this end, we used the gene-based proteome-wide association study (PWAS) method to detect associations mediated by the effects of variants on the protein function of all coding genes. The UK-Biobank (UKB) reports on 13,687 cases out of 274,824 participants of European ancestry, with a prevalence of 7.5% and 2.0% for females and males, respectively. The results from PWAS for ICD-10 E03 are a ranked list of 77 statistically significant genes (FDR-q-value <0.05) and an extended list of 95 genes with a weaker threshold (FDR-q-value <0.1). Validation was performed using the FinnGen Freeze 7 (Fz7) database across several GWAS with 33.5k to 44.5k cases. We validated 9 highly significant genes across the two independent cohorts. About 12% of the PWAS reported genes are strictly associated with a recessive inheritance model that is mostly overlooked by GWAS. Furthermore, PWAS performed by sex stratification identified 9 genes in males and 63 genes in females. However, resampling and statistical permutation tests confirmed that the genes involved in hypothyroidism are common to both sexes. Many of these genes function in the recognition and response of immune cells, with a strong signature of autoimmunity. Additional genetic association protocols, including PWAS, TWAS (transcriptional WAS), Open Targets (OT, unified GWAS) and coding-GWAS, revealed the complex etiology of hypothyroidism. Each association method highlights a different facet of the disease, including the developmental program of CH, autoimmunity, gene dysregulation, and sex-related gene enrichment. We conclude that genome association methods are complementary while each one reveals different aspects of hypothyroidism. Applying a multiple-protocol approach to complex diseases is expected to improve interpretability and clinical utility.

## Introduction

Hypothyroidism is a disorder of the endocrine system in which the thyroid gland does not produce enough hormones, or when the thyroid hormones act inadequately in target tissues ^1^. Hypothyroidism (EFO: 0004705) and its extreme condition, myxedema (EFO: 1001055), are signified by impairment in the function of the thyroid. The thyroid gland is crucial to the metabolism of all tissues and the early development of the central nervous system (CNS) ^2^. The majority of hypothyroidism cases are acquired due to iodine deficiency, with about 10% of the world population exhibiting some level of iodine deficiency ^3, 4^. In the USA, the manifestation of hypothyroidism is estimated to affect 1 in 300 people, but subclinical hypothyroidism occurs in 4–8% of the population ^5^. It is estimated that 1 in 8 people will develop functional deficiency of the thyroid in their lifetime ^6^.

The incidence rate of severe Congenital hypothyroidism (CH) is 1 in 4000 live births. Primary CH, which is associated with a missing or underdeveloped thyroid (dysgenesis), is the most common neonatal disease and accounts for most CH. While primary hypothyroidism is due to the failure of the thyroid gland itself, secondary disease is due to either transient or permanent dysfunction of the pituitary and its TRH or TSH signaling. In different parts of the world, the incidence of hypothyroidism increases with age and has a higher occurrence in females than in males ^7, 8, 9^. The diagnosis of hypothyroidism is based on blood tests measuring the levels of both free and bound thyroid hormones, TSH, and often also the presence of autoantibodies to thyroid markers ^10^. The majority of the cases can be assigned to hypothyroidism with autoimmunity (e.g., Hashimoto’s thyroiditis and autoimmune hypothyroidism (e.g., Hashimoto, Ord’s thyroiditis ^11^). CH is a monogenic developmental abnormality affecting the hypothalamic-pituitary-thyroid (HPT) axis ^12^. Unfortunately, the causal genetics of dysgenesis of the thyroid is limited to only 5% of the cases ^13^. Mutations in numerous thyroid transcription factors (TITF-1, TITF-2, PAX-8, FOXE1, GLIS3) are mostly syndromic ^14^. A systematic CH screen in Japanese ^15^ and Czech ^16^ confirms the challenge of identifying causal mutations. While most pathogenic variants of the TSH receptor (TSHR) are non-syndromic, mutated Gsα (GNAS1) and PDE8B, which are components of TSHR signaling, are linked with syndromic disease ^17^. Candidate genes that potentially disrupt thyroid gland formation have been linked to other rare monogenic diseases (reviewed in ^18 32^. Genes that cause dysfunction of thyroid hormone synthesis and secretion (dyshormonogenesis) act in the biosynthesis and cell biology of the thyroid hormones ^19^. Among these genes are thyroid peroxidase (TPO), thyroglobulin (TG), sodium iodide symporter (NIS), pendrin (PDS), thyroid oxidase 2 (THOX2), and iodotyrosine deiodinase (IYD). The iodothyronine transporter (MCT8) that is expressed in the thyroid gland membrane was also shown to drive hypothyroidism, which is coupled to neurological deficits ^20^. While most cases of CH occur sporadically, dyshormonogenetic cases are often recessively inherited ^21^. Importantly, CH is frequently associated with an increase in neonatal malformations, which can result in further disease complications ^22 34^. Interestingly, the occurrence of mutated CH causal genes differs substantially across populations ^23^.

The most common thyroid disorder in children is autoimmune thyroiditis (AIT) ^24^. AIT reflects some unknown defects in immunoregulation, which are translated into injury of the thyroid tissue, which in turn activates apoptotic cell death and thyroiditis. The genetic basis for AIT is unknown, but it is likely to combine genetics (estimated to account for 70% of the cases) and environmental factors that interact with the predisposed genetics. Autoantibodies against thyroid-specific antigens (e.g., TSHR, TG) were found in most patients ^24^. AIT belongs to a complex immune disorder where each gene contributes only a small effect, but tens of immune-susceptibility genes may collectively govern the immune recognition and immune response. Some of these genes were also identified in the thyroid autoimmune Graves’ disease (GD). Among these shared genes are HLA-class II (HLA-DR), Protein tyrosine phosphatase 22 (PTPN22), and Cytotoxic T lymphocyte antigen 4 (CTLA4) ^25^. Our understanding of the environmental factors that contribute to disease development is fragmented and mostly unknown. Risk factors may include hormones (e.g., estrogen), stress, smoking, and dietary iodine consumption. Importantly, hypothyroidism is linked to a higher incidence of other organ-specific autoimmune diseases ^24^.

In this study, we asked whether the genetic signature of ICD-10 of E03 in the adult population is dominated by CH dysgenesis and dyshormonogenesis, or by non-developmental genetics. We also ask whether the genetic effects of hypothyroidism/myxedema are associated with sex. To this end, we applied a gene-based method from the Proteome-wide association study (PWAS) that detects gene-phenotype associations through the effect of variants on protein function. PWAS aggregates the signal from all variants affecting each protein-coding gene with the assumption of dominant and recessive inheritance ^26^. We identified a gene-association predisposition and highlighted the prominent genetic signal effect in females that strongly overlaps with autoimmunity. This enabled a natural division of hypothyroidism into several subgroups that meet the definitions of congenital and acquired diseases. Our findings can improve our understanding of the role of the immunity signature in the biology of hypothyroidism in the population and suggest a sex-related approach for disease management.

## METHODS

### UKB processing

The UK Biobank (UKB) is a population-based database with detailed medical, genotyping, and lifestyle information covering ∼500k people at ages 40-69 across the UK who were recruited from 2006-2010. Analyses herein were based on the 2019 UKB release. We restricted the analysis to European origin [codes 1, 1001, 1002, 1003, respectively, in Ethnic background, UKB data field 21000]) and classified according to the genetic ancestry (Genetic ethnic group, data-field 22006). We further removed genetic relatives, by randomly keeping only one representative of each kinship group.

Disease classification is based on clinical information encoded by ICD-10 codes of hypothyroidism (E03) within main or secondary diagnosis codes (UKB data fields 41202 and 41204) and the summary 41270 covers the distinct diagnosis codes a participant has had recorded across all their hospital inpatient records in either the primary or secondary position. These fields cover ICD-10 from E03.0 to E03.9 (total of 29,478 participants), with 98.5% of them are marked as “unspecified hypothyroidism”, “other specified hypothyroidism” (E03.8, 0.7%), “hypothyroidism due to medicaments and other exogenous substances” (0.3%, E03.2) and CH (0.3%, E03.0-E03.1). This set of E03 includes 2,557 males and 11,094 females.

### ICD-10 E03 hypothyroidism genetic analysis

The UKB released genotyped data for all participants. The UKB genotyping scheme is based on ∼820,000 preselected genetic variations (from genotyping data of the UKB Axiome Array). Based on the UKB imputation protocol, the number of variants was expanded to the 97,013,422 imputation variants provided by the UKB. All together we tested all variants that are included in the 18,053 coding genes, including splicing variants. This set included 639,323 of the 97,013,422 of the whole genome imputed variants.

### Comparative and validation analyses

We used the Open Targets (OT) platform to select current knowledge on hypothyroidism ^27^. The Open OT Genetic platform (release date 3/2022) association score unifies evidence for drugs, text mining, animal models and genetic associations. A total of 1889 genes were listed. From the OT database, the list of mapped genetic variants to genes for hypothyroidism comprises 715 associated genes, from several large-scale independent GWAS, ranked by their summary statistics ^28^.

A unified OT global score (range 0-1.0) is determined by combining the gene-disease knowledge (e.g., literature, animal models, drugs) with the genetic association (GA) score. We extracted the SNPs associated with hypothyroidism and used OT genetic association scores to list the top variants for comparative analysis. Most informative associations were reported from several large studies including a hypothyroidism study from UKB in multiple ethnic groups ^3^, Neale’s lab study (2018, V2) focused on European ancestry with 361,141 participants among them 17,574 cases reported with “hypothyroidism/myxoedema (self-reported, non-cancer)”, and the compilation of UKB and Iceland (30,234 cases and 725,172 controls). Each gene is ranked by a genetic-association normalized scores (ranges 0-1.0). In addition, each gene is associated with an overall OT score that includes information from drug use, results from animal models and literature.

Validation was performed by comparing the gene association of PWAS and FinnGen-Fz7 (release date spring 2021), consisting of 309,154 individuals and supported by the Risteys R9 querying system. Several studies for PheWAS section 4 (Endocrine, nutritional and metabolic diseases, E4) were conducted in FinnGen.

### PWAS and functional effect scores

PWAS was developed as a gene-based association method and was shown to complement GWAS for a large collection of human diseases and human traits ^26^. PWAS methodology assumes that causal variants in coding regions affect phenotypes by altering the biochemical functions of the encoded protein of a gene. The functional impact rating at the molecular level (FIRM) pretrained machine-learning (ML) model, is then used to estimate the extent of the damage caused to each protein in the entire proteome ^29^. FIRM uses 1109 numerical features which are used by the ML model to predict variant-centric effect score (https://github.com/nadavbra/firm). FIRM performance was reported and validated for the pathological variants in ClinVar, reaching an AUC of 90% (precision = 86%, recall = 85.5%) and accuracy of 82.7% ^29^. Per-variant damage predictions are aggregated at the gene level from all listed variants (average of 35.4 nonsense and missense mutations per gene). A protein function effect score is reported for the dominant or recessive effects, as well as for a hybrid model of inheritance. PWAS analyzed gene effects for the UniProt-Swissprot (reviewed) human proteome. The current list of reviewed human proteins from UniProt includes ∼20k proteins, but unique and unambiguous mapping to Refseq gene names that were addressed by PWAS covers ∼18k proteins. Altogether, there are 2,119 reviewed proteins which are matched with gene names that are not included (Supplementary **Table S1**). Several of these missed genes are characterized by multiple protein versions and thus their gene to protein in undefined. Others, belong to endogenous retrovirus group (e.g., ERVK) or to constant and variable regions of immunoglobulins (e.g., IGLV. IGKV). These genes are not available as genuine genes in RefSeq.

The gene-based aggregation of the FIRM score associated with each variant allows the result of PWAS to cumulatively enhance weak signals associated with individual SNPs. In PWAS, each non-synonymous variant is assigned a functional effect score that aims to capture its propensity to damage the gene’s protein product, considering missense, nonsense, frameshift, in-frame indel, and canonical splice-site variants. The predicted effect score of a variant is a number between 0 (complete loss of function, LoF) and 1 (no functional effect). Variations other than non-synonymous (i.e., nonsense, frameshift, splice site variants) scored 0 for a complete LoF. We explicitly treated in-frame indels (see ^26^).

### Sex specific statistical analyses

We applied a procedure for assessing the possibility that the discovery of sex-specific PWAS significant results is due to the difference in the cohort sizes (2,557 males and 11,094 females). We applied random sampling of the sex-specific ICD-10 E03 cases according to the size of the males’ group. We performed 100 PWAS analyses (with repetitions) and identified the number of occurrences (up to 100 times) for males and females on the basis of equal group sizes.

We further performed a gene-based permutation analysis using the machine learning library extensions of python (mlxtend v0.21.0) in https://github.com/rasbt/mlxtend. We questioned whether genes were differentiated between sex-specific populations directly from the scores of the individual gene-level risk scores. Recall that the PWAS aggregated risk score per gene and per person is extracted from the calculated FIRM for the dominant and recessive scores. The analysis performed an association test between the risk score of each person and the label. Specifically, a permutation over the score of each gene, for testing whether the population of males that were diagnosed with ICD-10 E03 is distinct from that of the female population. We test the statistics of the results by running 100, 500, 1000, 5000, 10,000, 15,000, and 30,000 permutations.

### Comparable GWAS analysis

To perform an unbiased comparison between PWAS and GWAS for hypothyroidism, we used the same list of 172 covariates that includes sex (binary), year of birth (numeric), 40 principal components (PCs) that capture the ancestry stratification (numeric), the UKB genotyping batch (one-hot-encoding, 105 categories), and the UKB assessment centers associated with each sample (binary, 25 categories). We tested the 804,069 tagged genetic markers genotyped by the UKB and the imputed coding variants that included 639,323 of the 97,013,422 variants provided by the UKB. We tested the imputed variants that affect the coding region of 18,053 protein-coding genes. This is the identical set of variants (total 639,323) that PWAS considered. When testing raw genetic markers with GWAS, we used the convention of 0/1/2 variant encoding to specify the alternative allele. For the imputed variants we calculated the probabilistic expected number of alternative alleles ^26^.

### Statistical tests

For the effect size of a gene, we applied the statistical measure of the Cohen’s d value to measure the difference between two means divided by a standard deviation (SD) for the data, also known as standardized mean difference. For GWAS, the variant association and effect size were calculated by PLINK 2.0 default logistic regression, which produces the z score to specify effect size and its directionality. In GWAS, a positive z score indicates a positive correlation between hypothyroidism and the number of alternative alleles, thereby indicating a risk variant. Note that the effect-size metric results in PWAS and GWAS have different interpretations. In GWAS, a positive value indicates a risk variant. In PWAS, on the other hand, positive values indicate a positive correlation with the gene effect scores, whose higher values mean less functional damage, thereby indicating protective genes. Thus, negative values are indicative of protective variants in GWAS vs. risk genes in PWAS. We applied statistical tests by protocol for count down and a Spearman rank correlation test for testing the hypothesis that genes identified for males and females originated from identical probabilities.

## Results

### Comparative GWAS results for hypothyroidism

Large scale GWAS have been performed on several cohorts for hypothyroidism. A comparative study compiling six of the largest studies is shown in **Figure 1**. The comparison is performed with a root of the GWAS of “Hypothyroidism/myxoedema (non-cancer, self-reported)”, from Neale v2, 2018 study of the Europeans origin in UKB, covers ∼17.5k cases and ∼345k controls. There are 115 significant lead variants with p-value <5e-8. However, mapping a lead variant to a causal gene is nonconclusive and often inaccurate. Instead, each lead variant is associated with an expended credible gene list, which may include tens or hundreds of genes per independent locus. To focus on the most relevant variants and genes, we compared the results from additional 5 GWAS studies including the UKB Saige (2018) based on 28 million imputed variants (45 loci, p-value < 5e-8). The other GWAS (Supplementary **Table S2**) are based on large cohorts ranging from 210k to 580k participants with the number of cases ranged from 15k to 30k and the number of independent loci is 65 ^30^, 160 ^3^, 133 ^31^ and 50 from FinnGEN (Freeze 5, 2021) (**Supplementary Table S2)**. Unification of all the significant variants resulted with 20 variants that intersected all these six GWAS (**Table 1**).

**Figure 1.**
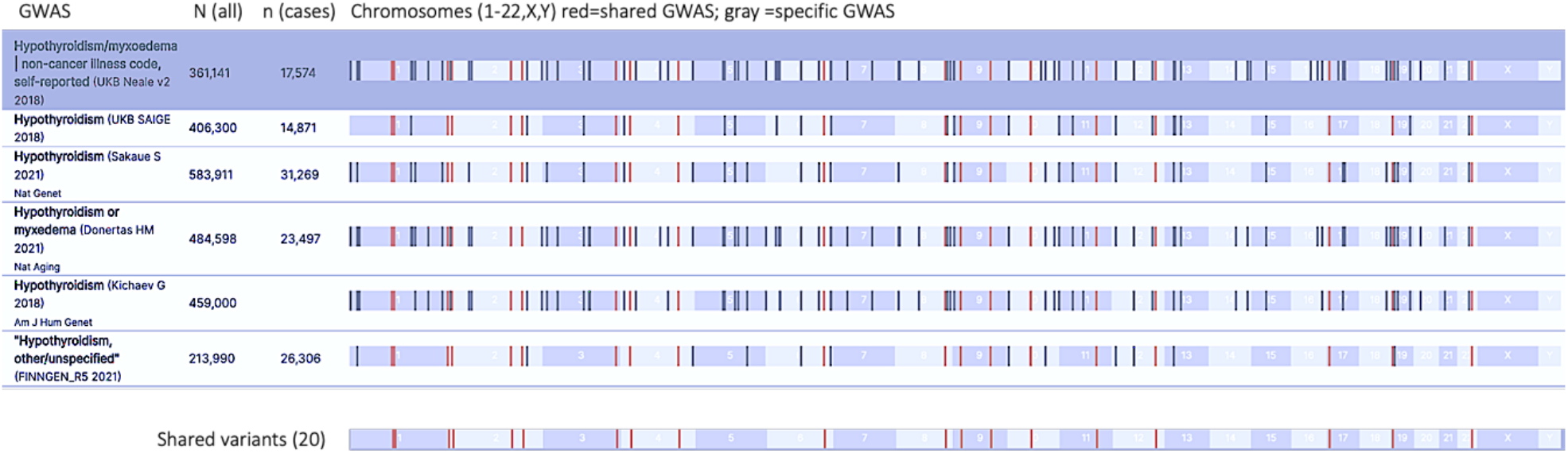
Summary of independent loci identified from major GWAS results as compiled in Open Target (OT) genetic portal. The total number of participants in each study is marked as N (all) and the n (cases) indicates the number of hypothyroidism cases. The positions mark in red font are those shared by all six studies. Note that some studies are based on overlapping cohorts. The positioning along the chromosomes (from left to right) of the shared 20 variants are shown below in red.

**Table 1.** The 20 shared variants from all available GWAS for hypothyroidism

| # | SNP rsID | Cytoband | Variant | Gene | AF<sup>a</sup> | # LD genes |
| --- | --- | --- | --- | --- | --- | --- |
| 1 | rs78765971 | 1p13 | 1_107819547_GAC_G | VAV3 | 0.09 | 3 |
| 2 | rs484959 | 1p13 | 1_109823461_T_C | GSTM3 | 0.5 | 23 |
| 3 | rs2476601 | 1p13 | 1_113834946_A_G | PTPN22 | 0.89 | 15 |
| 4 | rs11675342 | 2p25 | 2_1403856_C_T | TPO | 0.44 | 3 |
| 5 | rs1534430 | 2p24 | 2_12504610_C_T | TRIB2 | 0.41 | 1 |
| 6 | rs2111485 | 2q24 | 2_162254026_A_G | FAP | 0.62 | 6 |
| 7 | rs7582694 | 2q32 | 2_191105394_C_G | STAT4 | 0.78 | 7 |
| 8 | rs76897057 | 3q28 | 3_188407079_TA_T | LPP | 0.48 | 1 |
| 9 | rs34046593 | 4p15 | 4_26109971_G_A | RBPJ | 0.31 | 6 |
| 10 | rs546532456 | 4q31 | 4_148724495_C_CTT | NR3C3 | 0.0014 | 1 |
| 11 | rs9497965 | 6q24 | 6_148200156_C_T | SASH1 | 0.38 | 1 |
| 12 | rs2445610 | 8q24 | 8_127184843_A_G | POU5F1B | 0.35 | 2 |
| 13 | rs2123340 | 9p21 | 9_21589042_G_A | IFNE | 0.65 | 19 |
| 14 | rs7850258 | 9q22 | 9_97786731_A_G | FOXE1 | 0.66 | 14 |
| 15 | rs71508903 | 10q21 | 10_62020112_C_T | ARID5B | 0.18 | 4 |
| 16 | rs4409785 | 11q21 | 11_95578258_T_C | SESN3 | 0.17 | 5 |
| 17 | rs3184504 | 12q24 | 12_111446804_T_C | SH2B3 | 0.52 | 17 |
| 18 | rs61759532 | 17p13 | 17_7337072_C_T | ACAP1 | 0.23 | 59 |
| 19 | rs10424978 | 19p13 | 19_4837545_C_A | TICAM1 | 0.58 | 21 |
| 20 | rs229540 | 22q12 | 22_37195250_T_G | C1QTNF6 | 0.42 | 25 |

<sup>a</sup>AF, Allele frequency for non-Finnish European population.

The identified leading variants **(Figure 1)** are listed along with the most likely associated genes **(Table 1)**. For most variants, the allele frequency of non-Finnish Europeans is quite high, with the exception of 0.14% for rs546532456 (4q31). Note that for many variants, a large number of genes is reported and could not be separated due to strong genetic linkage, reflected by LD (linkage disequilibrium). In these cases, no conclusive assignment to a particular gene was possible, with only 4 of the 20 lead variants associated with a unique gene (**Table 1**).

As GWAS results are mostly driven by the statistics of the effect sizes, we expect that the shared variants from the multiple large-scale GWAS may explain variants signified by strong genetic effects, such as those occurring in congenital hypothyroidism (CH). Indeed, several of the listed genes are involved in thyroid hormone production and secretion. For example, the key enzyme thyroid peroxidase (TPO) acts in the iodination of tyrosine residues in thyroglobulin and thyroid hormones. An additional affected gene is the transcription factor FOXE1 that is implicated in thyroid gland morphogenesis (**Table 1)**. Both genes were implicated in CH by OMIM. TPO is reported as causal for Thyroid dyshormonogenesis 2. The relevance of FOXE1 was confirmed in the Chinese population ^32^, where abnormal expression of FOXE1 causes CH-based thyroid dysgenesis (OMIM 218700).

### PWAS results for hypothyroidism (ICD-10 E03)

The inherent limitation of GWAS in assigning a variant to a specific causal gene reduces the interpretability of the results. To overcome these limitations, we applied PWAS that exclusively focuses on the alteration in the coding region of the gene and assesses the impact of damaging variants on the protein function ^26^. Moreover, GWAS consider additivity (encoding as 0/1/2 for the alternative alleles) and thus identify mostly dominant heritability. A benefit of PWAS as a gene-based method is the ability to model a recessive and dominant mode of heritability (as well as hybrid mode that is based on having either heritability mode). Based on the UKB cohort for ICD-10 E03, we identified 77 statistically significant PWAS genes (FDR-q-value <0.05) and 95 genes with a weaker threshold (FDR-q-value <0.1). We then asked whether we could identify genes that are likely to be either recessive, dominant or hybrid inheritable models. Altogether, we identified 9 variants in genes (11.7%) that are recessive and 41 (53.1%) that are dominant. For the rest, the difference in the significant for between the models were <-log10(p-value) of 3.0 (**Figure 2A**). The fraction of recessive gene remains 12% also for the extended PWAS list (FDR <0.1; total 95 genes).

**Figure 2.**
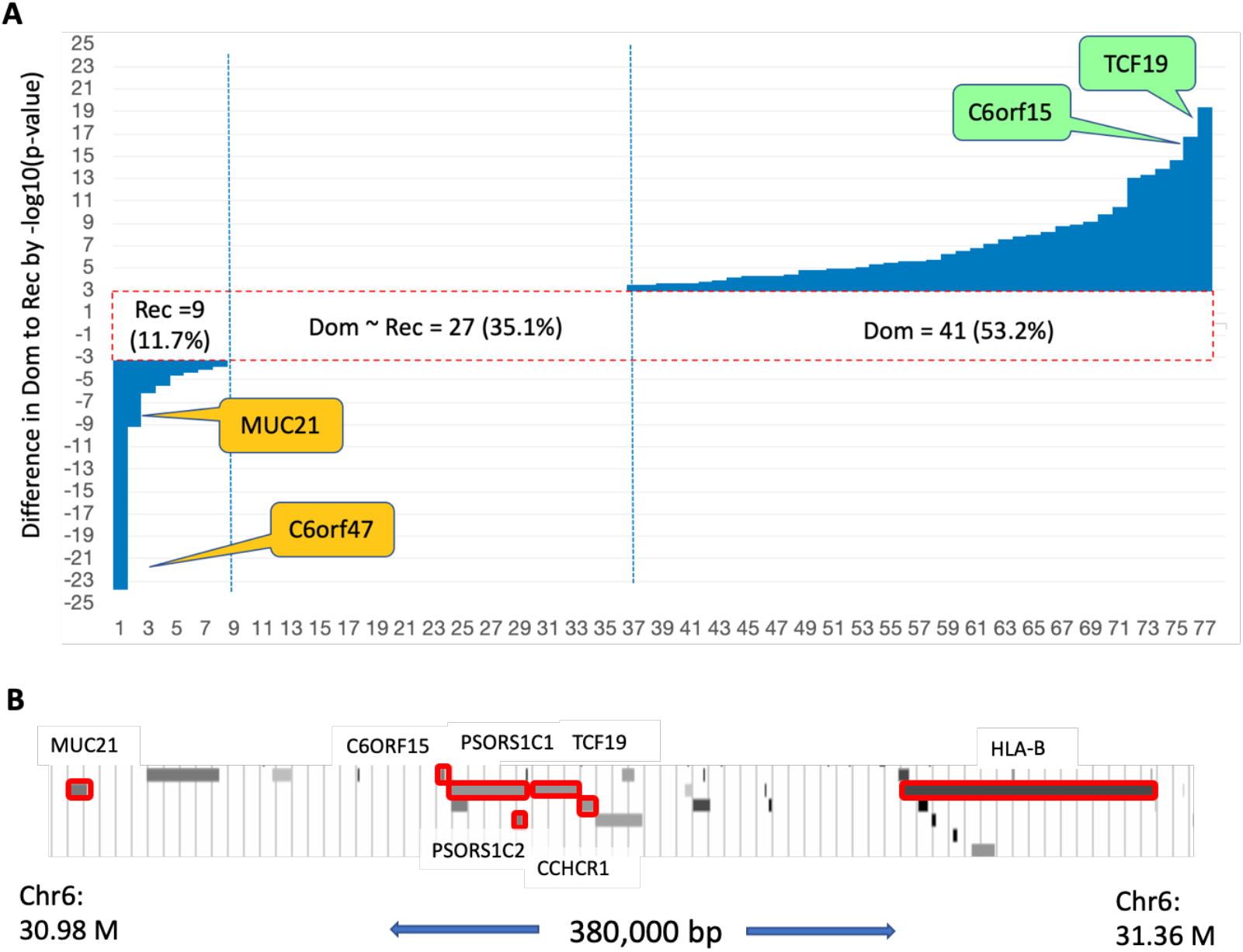
PWAS genes. **(A)** Difference in the statistical significance measured with a recessive and dominant inheritance model for each of the 77 identified genes (PWAS FDR <0.05; Rec and Dom, respectively). We set the difference in -log10(p-value) to be at least 3 to depict a heritability preference. The symbol of two most significant genes for the recessive (yellow) and dominant (green) models are indicated. **(B)** A 380 kbp region, ranging from MUC21 to HLA-B, is shown. In this restricted locus, PWAS identified 7 genes (red frames).

The genes ranked highest by the recessive heritability signal are C6orf47 and MUC21. While C6orf47 has no functional information, MUC21 belong to class I major histocompatibility (MHC) region. Both genes are located in the same chromosomal locus (6p21.33, at a distance of 0.66M bp). The recessive signals associated with PSPH and HLA-DPB1 are substantial but the signal for the rest of the others recessive genes (CD86, PCSK7, C1QTNF6, FAM221B and CTLA4) is only modest. The genes that carry the strongest dominant inheritance signal include TCF19 (transcription factor SC1) and C6orf15. Both genes are located at the same locus (6p21.33 at a distance of <200 kbp). We observed that many of the genes identified belong to the highly polymorphic region of the major histocompatibility complex (MHC). The MHC is a segment of ∼3.6M bp (chr6: 29.6M – 33.4M) with over 300 genes, about half of which are non-coding genes or pseudogenes. Many of the 141 coding genes in this region are cell surface proteins essential for the adaptive immune system. **Figure 2B** shows a small section of the MHC locus (380 kbp) including 7 of the PWAS identified genes.

We then analyzed the 77 significant genes by the effect size with respect to their heritability models (**Figure 3**). The PWAS most significant genes are listed along with their p-values FDR (q-value) and colored according to effect size, being risk increase or decrease for E03. Among the top ranged genes (FDR q-value, <1e-07; 26 genes, **Figure 3A)**, 80% are associated with an increased risk for hypothyroidism and only a few of the genes are associated with a reduced risk for this condition.

**Figure 3.**
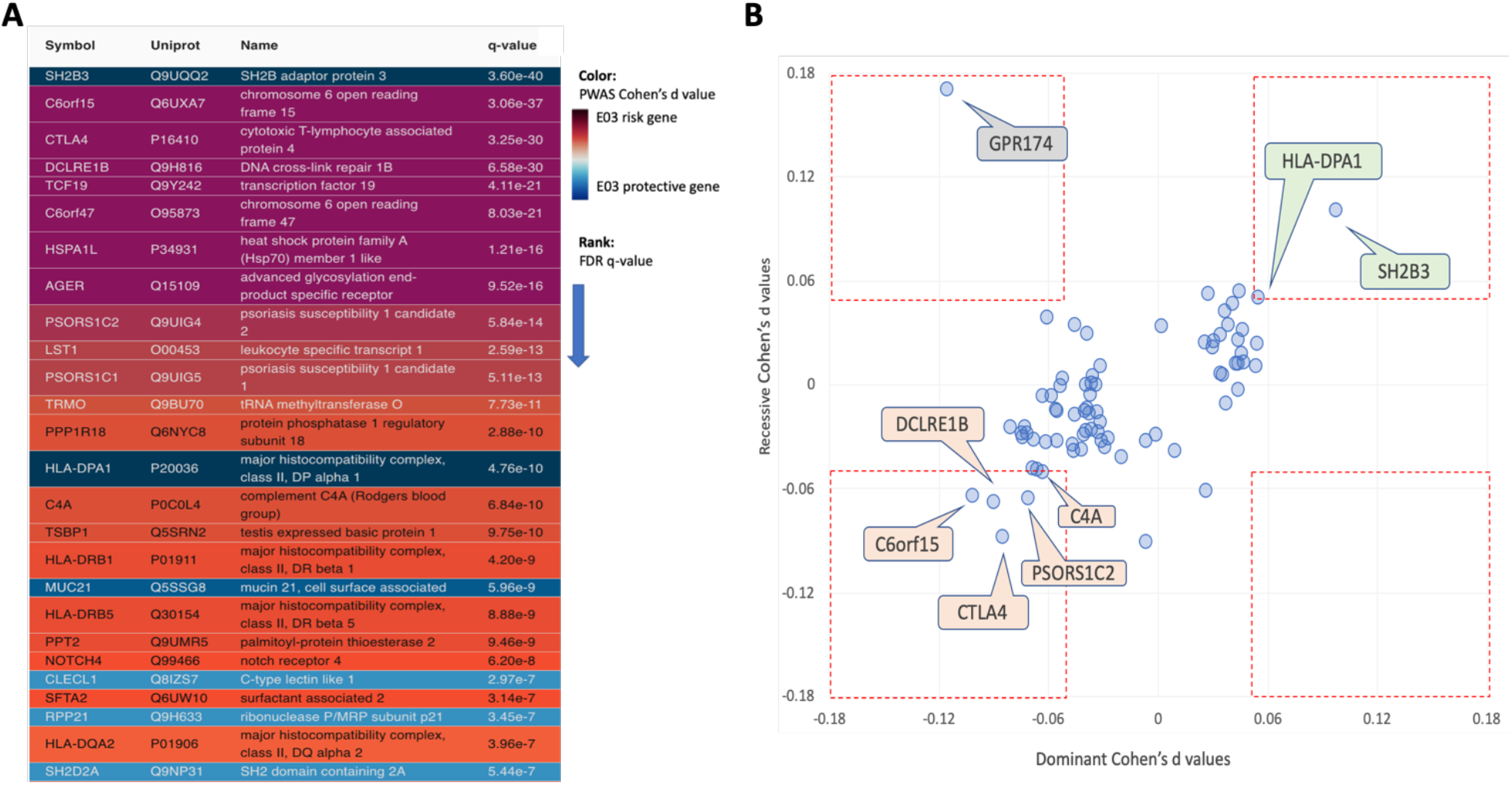
The associated risk for PWAS gene list by heritability model. **(A)** Top 26 highest statistically significant genes from PWAS for ICD-10 E03 list (77 genes) with q-value <1e-07. Genes with an increased and decreased risk are colored purple/red and blue, respectively. **(B)** Effect size (Cohen’s d) for PWAS results for the recessive and dominant models. The genes within the red frames are associated with Cohen’s d below an absolute value of 0.05. Positive Cohen’s d values are associated with a reduced risk. The 8 genes with absolute effect size smaller than -0.05 or larger than 0.05 are labelled. Genes with protective effects are colored green and risk genes as light red. **Supplementary Table S3** lists all genes and their statistics.

We observed that most genes have rather limited effects (absolute values are smaller than 0.05, **Figure 3B**). For the dominant model, 23 genes exhibit Cohen’s d values lower than -0.05, indicative of an elevated risk for hypothyroidism, with additional 4 genes are associated with a protective effect (Cohen’s d value >0.05). For the recessive model, we identified 7 genes with Cohen’s d values lower than -0.05, and 5 genes that are likely to have a protective effect (i.e., Cohen’s d values > 0.05). Only GPR174 out of the 77 PWAS significant genes, displayed a conflicting heritability trend **(Figure 3B)**. GPR174 displays a strong protective effect for the recessive mode, but an increased risk in the dominant model. All other genes (**Figure 3B**, dashed red frames) are consistent with their heritability trend with two protective genes (SH2B3 and HLA-DPA1) and 5 genes that exert an elevated risk.

### Duplication and validation of hypothyroidism PWAS results

To test the relevance of the PWAS identified genes we searched for independent evidence from participants external to the UKB. To this end, we investigated the Finnish biobank (FinnGen) as an independent validation resource. We searched for the genes listed by the FinnGen for “Hypothyroidism, other/unspecified” (phenotype a, **Table 2**). The analysis is based on a recent version of FinnGen (Freeze 7.0, Fz7) with 38,554 hypothyroidism cases and 263,704 controls. Altogether there are 103 associated genes that were identified for 146 lead variants (some genes were repeatedly identified by independent lead variants and other failed to assign to a gene). Altogether, we validated 9 genes that overlap between the PWAS and FinnGen results. These genes are, listed according to their chromosomal location order, DCLRE1B, CTLA4, TLR3, HLA-DPB1, TRMO, PCSK7, SH2B3, THOC5 and C1QTNF6. **Figure 4** shows the Manhattan plot for hypothyroidism, other/unspecified by FinnGen and the PWAS validated genes. Several prominent genes that were identified in FinnGen were not included in the PWAS analysis. We marked several of such genes PTPN22, SYN2, TSHR, HSP89 by their chromosomal location in Chromosome 1, 3, 14 and 17, respectively.

**Table 2.**
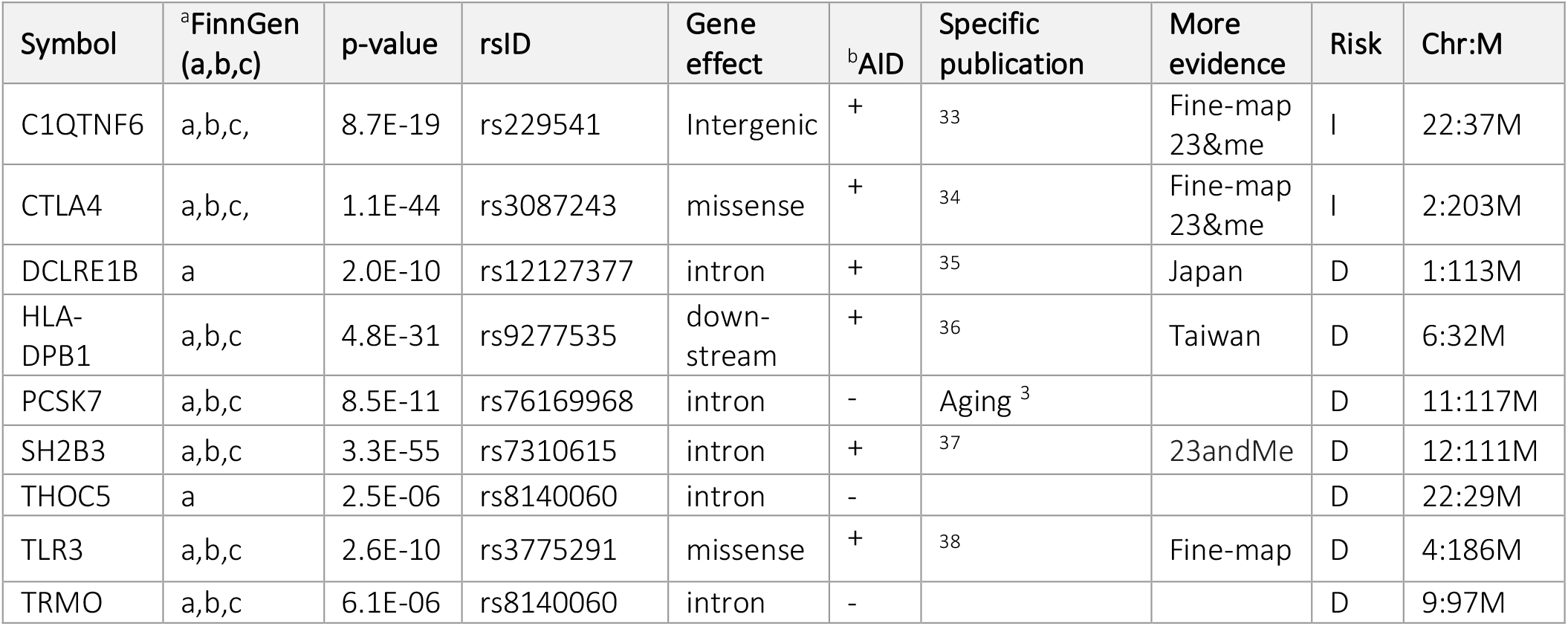
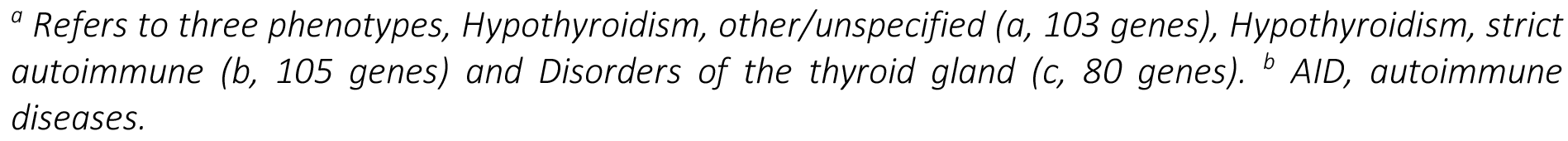
Validation of PWAS gene by FinnGen Fz7 for Hypothyroidism phenotypes

| Symbol | <sup>a</sup>FinnGen (a,b,c) | p-value | rsID | Gene effect | <sup>b</sup>AID | Specific publication | More evidence | Risk | Chr:M |
| --- | --- | --- | --- | --- | --- | --- | --- | --- | --- |
| C1QTNF6 | a,b,c, | 8.7E-19 | rs229541 | Intergenic | + | <sup>33</sup> | Fine-map 23&me | I | 22:37M |
| CTLA4 | a,b,c, | 1.1E-44 | rs3087243 | missense | + | <sup>34</sup> | Fine-map 23&me | I | 2:203M |
| DCLRE1B | a | 2.0E-10 | rs12127377 | intron | + | <sup>35</sup> | Japan | D | 1:113M |
| HLA-DPB1 | a,b,c | 4.8E-31 | rs9277535 | down-stream | + | <sup>36</sup> | Taiwan | D | 6:32M |
| PCSK7 | a,b,c | 8.5E-11 | rs76169968 | intron | - | Aging<sup>3</sup> |  | D | 11:117M |
| SH2B3 | a,b,c | 3.3E-55 | rs7310615 | intron | + | <sup>37</sup> | 23andMe | D | 12:111M |
| THOC5 | a | 2.5E-06 | rs8140060 | intron | - |  |  | D | 22:29M |
| TLR3 | a,b,c | 2.6E-10 | rs3775291 | missense | + | <sup>38</sup> | Fine-map | D | 4:186M |
| TRMO | a,b,c | 6.1E-06 | rs8140060 | intron | - |  |  | D | 9:97M |

<sup>a</sup> Refers to three phenotypes, Hypothyroidism, other/unspecified (a, 103 genes), Hypothyroidism, strict autoimmune (b, 105 genes) and Disorders of the thyroid gland (c, 80 genes). <sup>b</sup> AID, autoimmune diseases.

**Figure 4.**
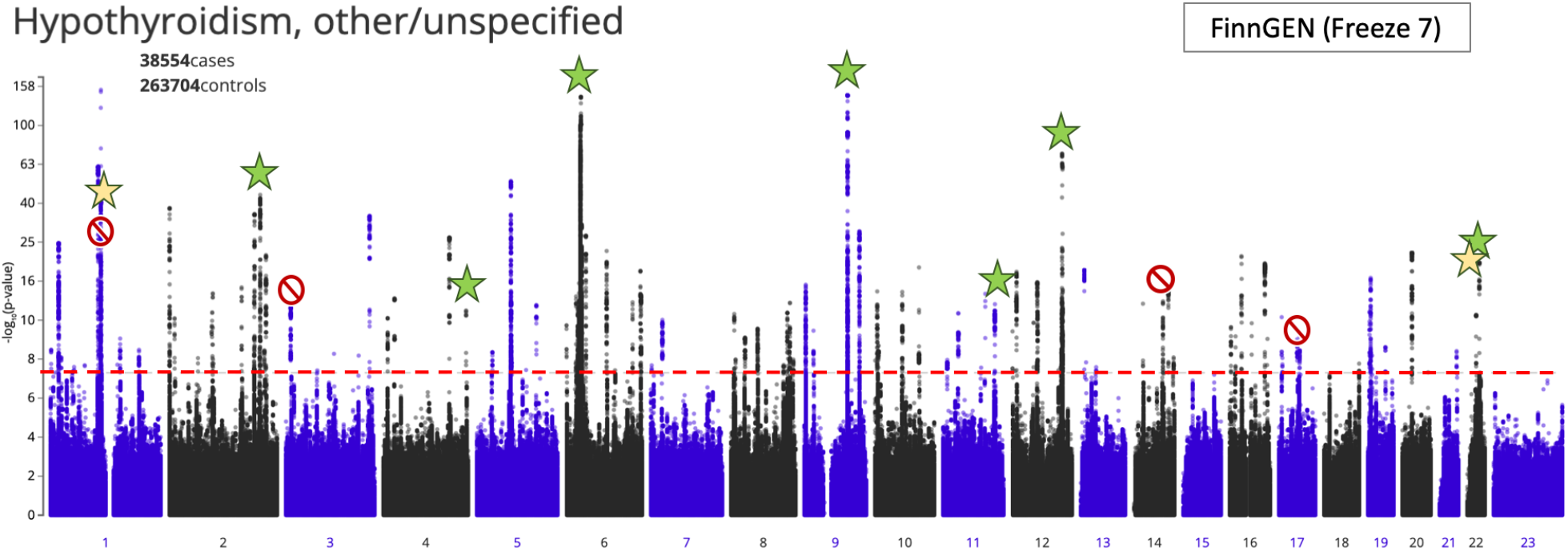
GWAS results for hypothyroidism, other/unspecific. The validated genes are marked (indicated by a star). Yellow stars indicate 2 genes that were validated by hypothyroidism, other/unspecific but not by the related phenotype of “Hypothyroidism, strict autoimmune” and “Disorders of the thyroid gland”. Green stars indicate genes that were validated in all hypothyroidism related phenotypes (7 genes). The 9 validated genes, ordered by chromosomal locus, are DCLRE1B, CTLA4, TLR3, HLA-DPB1, TRMO, PCSK7, SH2B3, THOC5, and C1QTNF6. Another 4 genes that are not included in our PWAS analysis are marked with a red symbol (PTPN22, SYN2, TSHR, HSP89). The horizontal line marks the PWAS threshold of p-value 5e-08.

**Table 2** lists the 9 genes that are validated from the FinnGen data and shared with the PWAS discovery. Testing the stability of the analyses, we tested GWAS for related phenotypes. We reanalyzed the narrower phenotype of “Hypothyroidism, strict autoimmune” (33.4k cases, 227.4k controls, phenotype b), and a more general term of “Disorders of the thyroid gland” (45.5k cases, 263.7k controls, phenotype c). The overlap with respect to “Hypothyroidism, other/unspecified” with these two phenotypes was 86.05% and 84.07%, respectively. The number of unique genes was 105 and 80 for the “Hypothyroidism, strict autoimmune” and “Disorders of the thyroid gland” (refers to as phenotypes b and c), respectively. A further validation of significant genes is based on an independent cohort of 23&me ^11^. Several of the duplicated genes were supported by fine-mapping, an example of which is shown in Supplementary **Figure S1** for the TLR3 gene.

The overlap between PWAS discovery and FinnGen gene list is statistically significant according to the hypergeometric probability test for FinnGen Hypothyroidism phenotypes (a, p-value: 6.4e-10; b, 2.6e-07; c 4.5e-08). **Table 2** lists the 9 validated genes (out of 77) along with the supported evidence. Many of the validated genes are associated with autoimmunity. For example, variants in C1QTNF6 on Chr22 are associated with Crohn’s disease and type 1 diabetes (T1D) (Supplementary **Table S3**). CTLA4 (cytotoxic T-lymphocyte associated protein 4) belongs to immunoglobulin (Ig) superfamily and the protein leads to inhibitory signal to T cells. Mutations in this gene have been associated with T1D, celiac disease (CD), systemic lupus erythematosus (SLE), Graves’ disease (GD), Hashimoto thyroiditis, and other autoimmune diseases. Both proteins are associated with an increased risk with odds ratio of ∼1.1. Interestingly, the HLA-DPB1 gene, which participates in almost all autoimmune diseases, is associated with a reduced risk for hypothyroidism but a substantial risk for other autoimmune diseases.

### Autoimmunity associated genes are enriched in PWAS results for hypothyroidism

Inspecting all identified genes by PWAS showed that 36 genes (47%) are located at the Chr6p22.1-p21.32 locus that specifies the MHC locus (hypergeometric p-value 7.3e-57). Many of these genes are associated with the immune system and, more specifically, with autoimmunity. It is known that autoimmune diseases are often caused by dysregulation of the acquired immune system. Thus, we asked whether the identified genes and their effect directionality may shed light on the underlying mechanisms for hypothyroidism. To this end, we reconstructed a connectivity map among the 77 PWAS genes as represented by STRING ^39^ (score >0.9, **Figure 5**). We found that 21 of the PWAS identified genes are functionally connected. This connectivity is highly significant (p-value of 3.67e-10). By lowering the STRING functionality threshold, more genes were included in the graph (i.e., 30 connected genes with a STRING Score >0.7). The network is mostly associated with cellular immunity, including antigen presentation, processing, and T-cell regulation. Relationships between MHC variants involved in autoimmunity determine other aspects of immunity, such as response to infectious diseases and inflammation. The strong connectivity observed (**Figure 5**) and the extreme enrichment in coding genes identified within the MHC locus (enrichment of >60 fold higher than expected) argues for a dominant genetic signal that combines hypothyroidism and autoimmune complex diseases ^40^. The HLA is extended to the membrane stimulation subnetwork through CD86 which is expressed on dendritic cells, macrophages, B-cells, and other antigen-presenting cells. CD86 provides the signal necessary for T cell activation and survival, the binding of CTLA4 to CD68 in an inhibitory switch to T-cell activation. In addition, the T-cell surface marker CD2 is among the hyperthyroidism PWAS identified list. GPR174 (G-protein-coupled receptor 174) is an orphan X-linked gene that is not part of the functional network (**Figure 5**). It displays a strong effect size (Cohen’s d value of 0.171) and was associated with autoimmune polyendocrine syndrome (APS). Interestingly, testing the missense mutations demonstrated a significant association with autoimmune Addison’s disease (AAD) with an odds ratio (OR) of 1.80 ^41^. Similarly, the non-synonymous GPR174 SNP rs3827440, resulted in a significant increased risk towards Graves’ disease (GD) ^42^. We confirm that the signal observed for the immunological signature is extended beyond the MHC loci.

**Figure 5.**
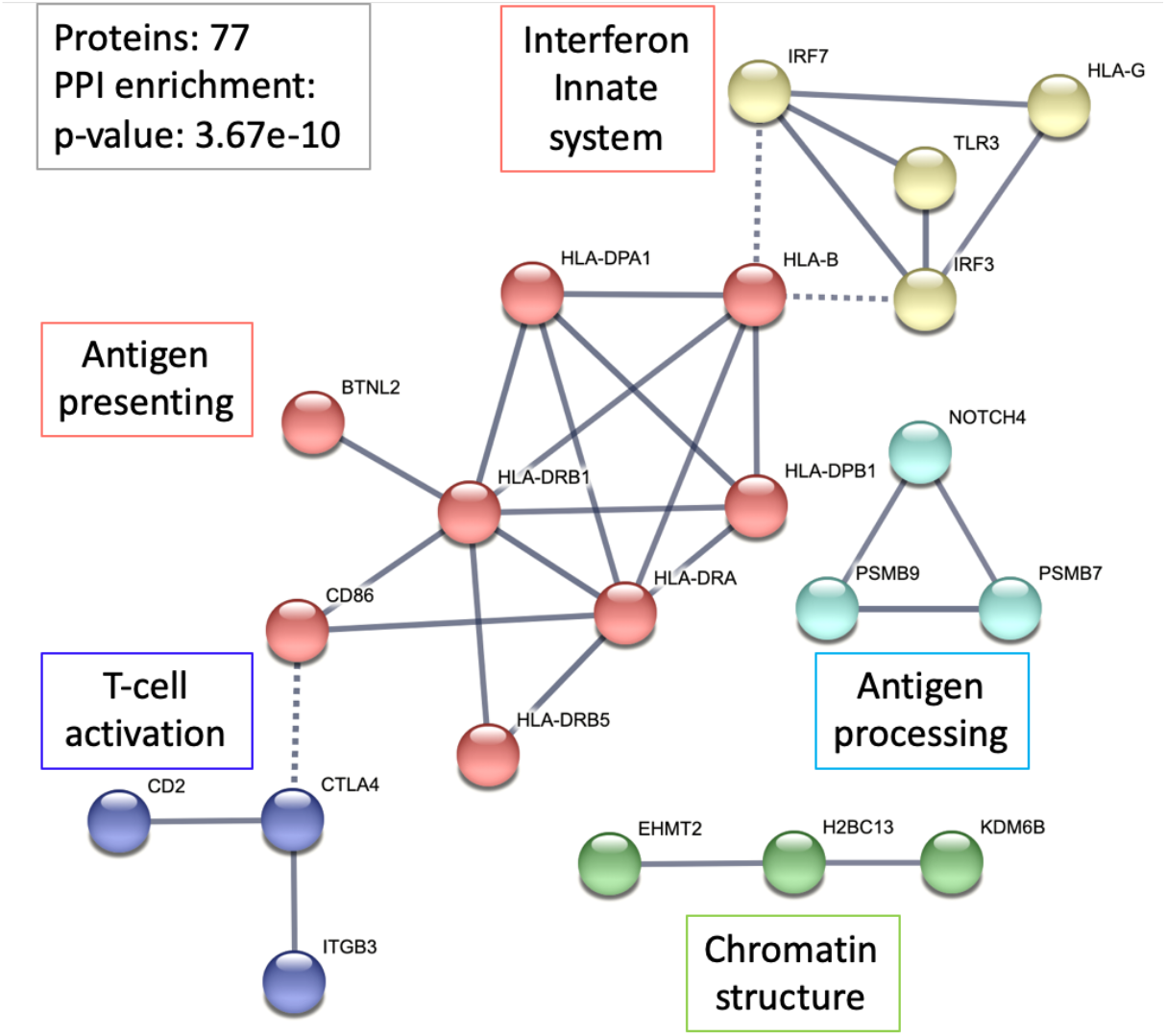
A network relation of the 77 proteins identified as significant by PWAS approach. The STRING network represents the genes connected at an interaction score >0.9. Specifically, the MHC locus is strongly represented (HLA-DPA1, HLA-DRB1, HLA-DRB5, HLA-B, HLA-DPA1, HLA-G). Dashed lines mark connection between clusters. A unified function for each cluster is indicated and colored (e.g., Antigen processing).

### Gene-centric association studies exposing functional complementarity in hypothyroidism

The analysis in **Figure 1** is based on GWAS as compiled in Open Targets (OT). The OT platform provides a knowledge-based resource that converts association of genes to diseases by including rich biological knowledge (e.g., from literature, animal models, pathways, drugs).

Altogether, OT reports on 1,889 hypothyroidism gene targets, of which 715 genes were scored by genetic association (GA score; range 0-1.0; **Figure 6A**, Supplementary **Table S4)**. We compared the PWAS results with the OT list based on genetic association. We found that the overlap of the lists is statistically significant (hypergeometric test p-value 6.38e-25; 10.5-fold enrichment). An even higher enrichment is found for genes selected by their GA score (total 222 genes >0.3, p-value 5.28e-13, 14.7-fold enrichment; **Figure 6A**). The observation that 58% of the PWAS genes are unique and not identified by the GWAS results reported by OT, raised the possibility that each association methodology exposes a different aspect of hypothyroidism. In testing this proposal, we analyzed the top ranked genes for hypothyroidism according to the OT global score of >0.5 (see Methods) and observed that none of the 25 genes found were identified as significant genes by PWAS. To gain insight on the relevance of these genes to hypothyroidism, we compared this selected OT list to GWAS results for congenital hypothyroidism (CH; supported by Orphanet (ORPHA:442; 43 genes). Remarkably, 24 of the 25 OT-reported genes scored >0.5, were included in the CH exclusive gene set. Moreover, these genes are functionally connected, whereas only SLC16A2 (**Figure 6B**) lacks direct genomic evidence for CH. We conclude that PWAS that extracts knowledge from the UKB population fails to identify causal CH genes. Instead, genes identified by PWAS are likely to contribute to acquired chronic hypothyroidism, and to the genetic signal of autoimmunity.

**Figure 6.**
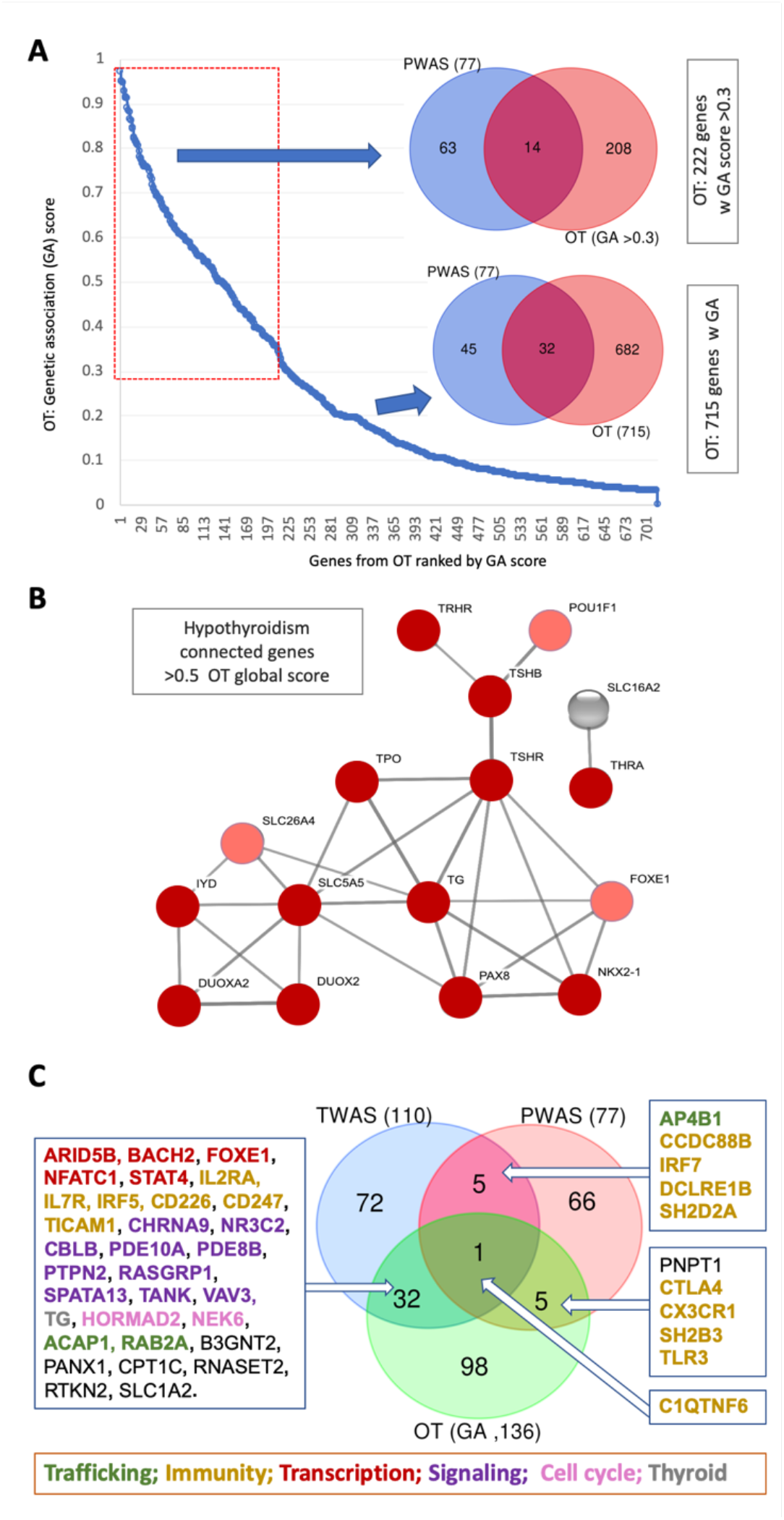
Genomic association by Open Targets (OT). **(A)** Ranked genes by their genetic association scores (total 715 genes). The overlap of 77 PWAS genes and the 222 OT genes with GA scores >0.3 and for all 715 genes. **(B)** A network relation of 25 OT genes that are ranked by the OT global score >0.5. The interaction of the top 25 genes resulted in a highly connected graph by STRING (edges are with >0.7 interacting confidence score). The nodes are colored according to the scores (dark red >0.5, light red <0.5). Only one protein lacks genetic association with CH. **(C)** Venn diagram of major association resources: PWAS (77 genes), GWAS (OT, by genetic association score >0.5 (136 genes) and TWAS, a transcription-based association (110 genes). The subsets of overlapping genes are marked according to their functional annotations.

To further substantiate the complementarity of the different association methods, we compared the PWAS gene set with genes resulted from transcriptome-wide association study (TWAS) ^43^. **Figure 6C** emphasizes the overlaps of association studies for hypothyroidism that relied on UKB entries of European origin: the PWAS (77), the GWAS from genetic association score >0.5 (136), and the TWAS significant expression-trait associations (110). The results show that the overlap between PWAS to OT or TWAS to OT is limited to 6 genes only, with the overlapping genes being exclusively related to cellular immunity. On the other hand, the overlap between TWAS and OT is higher and with 32 genes accounts for 25-30% of the gene sets.

The only gene that is shared by all three tested association studies in C1QTNF6 (C1q and TNF related 6). C1QTNF6 is known to carry two coding mutations, rs229527 (22:37,185,445:C:A) and rs229526 (22:37,185,382:G:C), that are associated with all types of hypothyroidism phenotypes (Supplementary **Figure S4**). Importantly, these variants are also associated with numerous autoimmune diseases including T1D, medication for Crohn’s disease, and more. In all cases, these variants increase the risk for the reported phenotype by a factor of 1.06-1.10. These TWAS-OT common genes act in TSH signaling, trafficking, and developmental processes, whereas the immunological genetic signal is less prominent. We conclude that comparison results from multiple association methodologies exposed many of the genes that specify CH and thyroid function.

### Gene-based association studies by sex

The higher prevalence of ICD-10 E03 in females relative to males raised the question whether hypothyroidism is signified by sex-dependent genetics. To this end, we applied PWAS gene-aggregative approach separately for males and females **(Figure 7**). The Venn diagram emphasizes that females displayed 63 significant genes while only 9 were reported for males. Moreover, out of the 63 PWAS significant genes, 7 genes (ARID3C, ATL2, HDGF, IRAK1, LRRC47, OR13A1 and SLC15A2) do not appear as significant in the full cohort combining both sexes (**Figure 7A**). Notably, HDGF (Heparin binding growth factor) resulted in a significant p-value (7e-08) in FinnGen “Hypothyroidism, other/unspecified”. In addition to its established role in cancer development, it was implicated in autoimmunity and specifically in rheumatoid arthritis (RA) ^44^.

**Figure 7.**
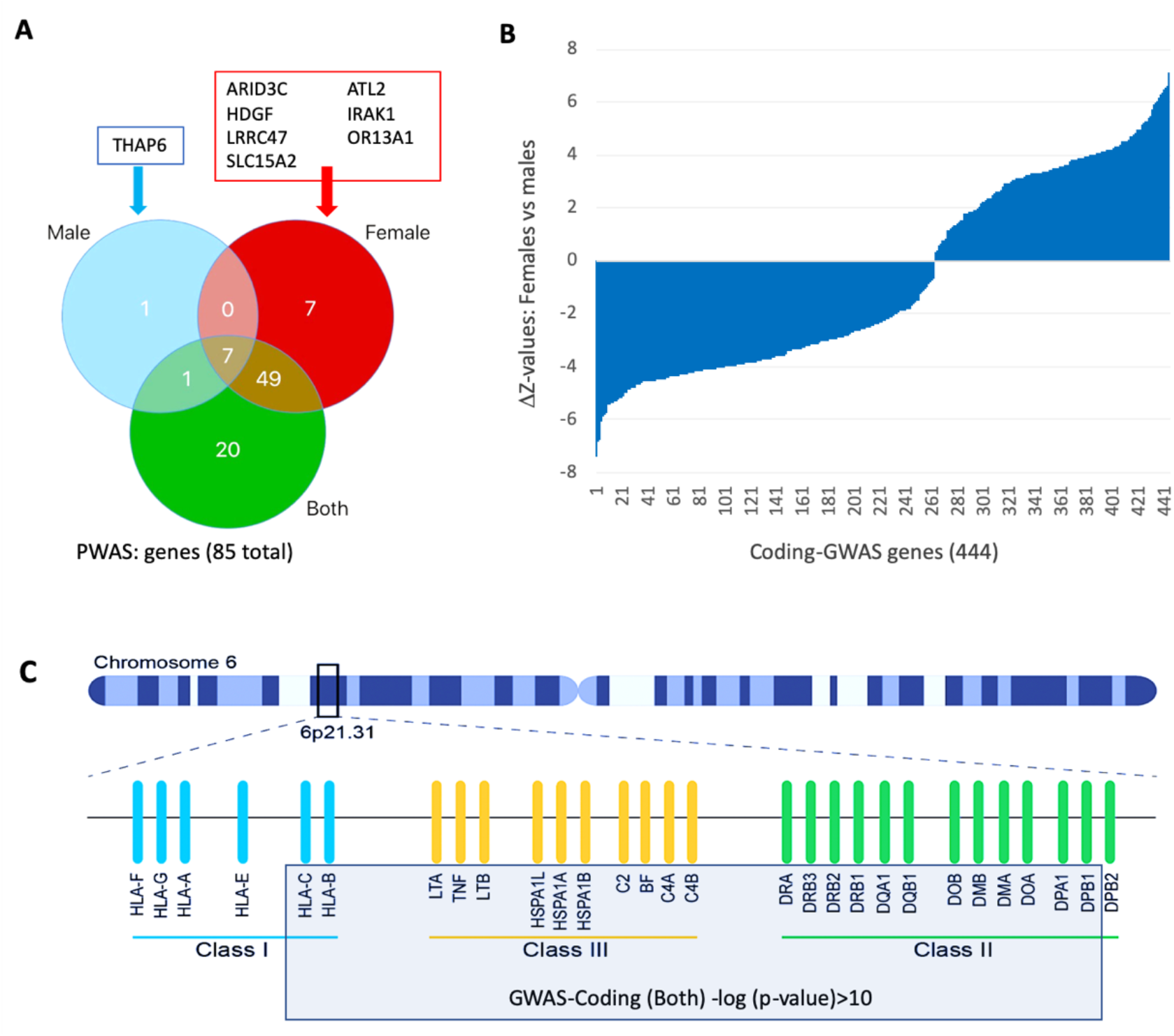
PWAS analysis by sex. **(A)** Venn diagram for the number of genes that are significant (q-value <0.05) by sex. Female and male exclusive genes are indicated. **(B)** GWAS-coding analysis covering 640k imputed variants and sorted by the difference in the z-values for females to males. Altogether there are 444 variants with -log10(p-value)>4.0 for either females or males. **(C)** Schematic view on the MHC-region on chromosome 6. Representative genes are indicated. There are 115 variants with -log10(p-value)>10 for both sexes, 93% are positioned in 2.4 Mbp from the MHC region (gray frame). The significant variants by sex are listed in Supplementary **Table S6**.

The genes identified in males are either shared by females (7 genes) or included in PWAS results from both sexes, with only one gene (THAP6, THAP domain containing 6) male-exclusive. Spearman rank correlation for the shared genes in females and males confirmed the high correlation and the similarity in gene effects between the sexes (correlation: 0.61, p-value 0.14). To minimize the effect of the group sizes of males and females, we randomly sampled among E03 females a subset that matched the smaller male group size (n=2,557) and performed 100 independent runs of PWAS with balanced group sizes. Altogether, 217 significant genes were collected, 70% of them were identified only by a single PWAS run (out of 100), and should be considered as noise. Identified genes, sorted by the number of occurrences across the 100 PWAS runs are listed in Supplementary **Table S5**. Genes that were detected in >20% of the runs show stable appearance and no difference between the sexes. The most statistically significant genes remain at the top of the list when the full size PWAS for each sex group was conducted (Supplementary **Figure S2**). We conclude that the reported difference in discovery rate for males and females is mostly attributed to group size differences.

We also performed permutation tests over the score of each gene (see Methods) to assess whether any of the identified gene directly display significant difference in their assignment to females of males following 100 to 30,000 such tests. In all cases we observed that the genes do not exhibit sex-dependent differences (Supplementary **Figure S2**), and already at 500 permutations the results are very stable. Only two genes consistently and significantly failed the permutation tests (p-value of 3.3e-05; Supplementary **Figure S2C**). One of these genes is IRAK1 (Interleukin-1 receptor-associated kinase 1) that is fundamental in initiating the innate immune response against pathogens. The IRAK1 participates in the cascade that drives cells in an antiviral state. The other gene, GPR174 (Probable G-protein coupled receptor 174) exhibits a strong effect size and conflicting heritability mode (**Figure 3B**). GPR174 is positioned on the X-chromosome and its impact on autoimmune Addison’s disease (AAD) and Graves’ disease (GD), and its female preponderance was confirmed ^41^. We conclude that all other identified genes do not show sex difference mechanistic differences between the sexes for hypothyroidism (Supplementary **Figure S2C**).

To further extend the view on hypothyroidism genetics stratified by sex, we performed a non-standard GWAS, coined coding-GWAS, that analyzes only coding and splicing variants which are identical to the set of variants used for PWAS (∼640k variants). The results are sorted by the difference in the statistical significance by sex using z-values (**Figure 7B**). We found that there are 444 variants where the difference in z-value is at least 4.0. There are 100 and 62 such variants for females and males, respectively. We found that the majority of the GWAS-coding variants were confined to the MHC-region on chromosome 6. **Figure 7C** shows a scheme of the MHC-region (3.6 Mbp) on chromosome 6. Out of the 115 variants with -log10(p-value)>10 for both sexes, 93% are positioned within a chromosomal segment spans 2.4 Mbp of the MHC region (gray frame). The list of 106 significant variants within the MHC region affected 28 genes. Many of these variants occurs in HLA-C, HLA-DRB1, HLA-DRB5 and HLA-DPA1.

### Gene-based polygenic risk estimates for hypothyroidism

The use of a polygenic risk score (PRS) for hypothyroidism can benefit personalized medicine by highlighting the subset of the population at risk according to the genetic landscape of the individuals ^11,31^. PRS for hypothyroidism (phenotype code HC643) from the European ancestry entries in UKB was calculated and showed a limited clinical utility ^45^. Specifically, the predicting performance for hold-out tests showed 0.151 and 0.072 using Tjur’s and Nagelkerke’s pseudo-R^2^ tests, respectively. Similar results were calculated for Hypothyroidism/myxedema (phenotype code HC219, Supplementary **Figure S3**). We performed a gene-based PRS approach based on PWAS for hypothyroidism (ICD-10, E03). Significant genes were analyzed according to heritability model (dominant or recessive considerations) and sex. In our approach, we examined the distribution of PWAS effect scores by considering all possible cutoffs for all subsets with effect scores that are either below or above a certain threshold. We calculated the UKB prevalence of 7.45% and 2.04% for females and males, respectively.

We demonstrate a gene-specific risk for SH2B3 (SH2B adaptor protein 3) across all 50 coding variants along the coding gene (**Figure 8A**) and indicated the variants that display stronger statistics as independent variants (p-values <0.05, **Figure 8B**). SH2B3 acts as a protective gene (q-value 1.81e-40). The histogram shows the PRS-like cohort partition relative to the overall prevalence (4.98%).

**Figure 8.**
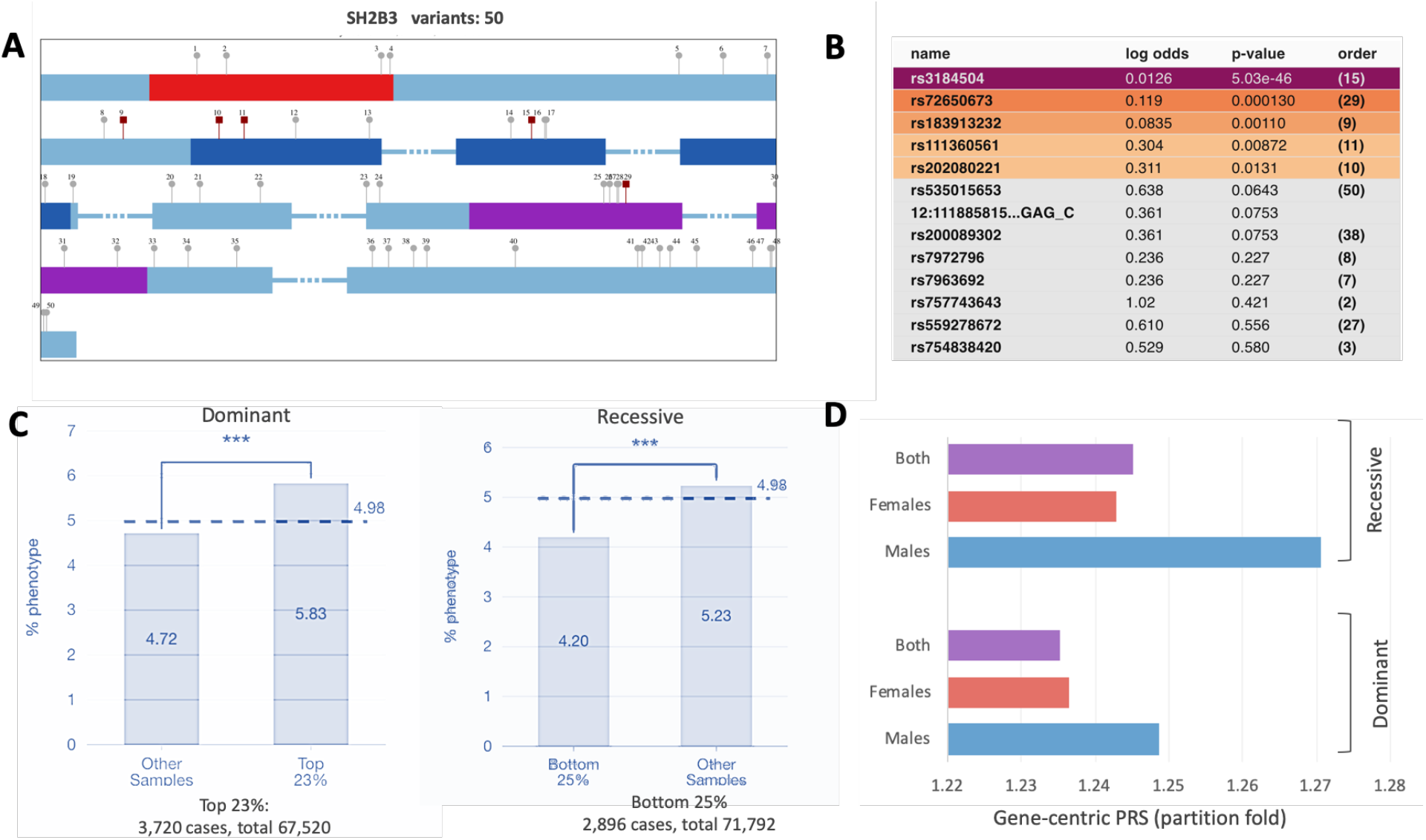
Dominant and recessive PWAS gene effects in gene-specific PRS-like statistics. **(A)** Schematic view of SH2B3 along with the Pfam domains: Phe_ZIP, PH and SH2, colored red, dark blue and purple rectangles, respectively and the 50 variants that are within the coding regions (UKB imputed). The lollipop symbols are indexed 1-50. These are the variants used to calculate the PWAS gene effect. The variants that are identified to be significant at p-value <0.05 are indicated by red square. **(B)** The list of significant variants, along with their statistics and calculated log odds. Colored are the GWAS variants with p-value <0.05, indicated as variants colored red in A. The index of the variants (1-50) is in parenthesis as in (A). **(C)** The plots show the most significant partitioning of individuals selected by the cutoff with the lowest p-value by Fisher’s exact test. The number of individuals for the best partition is shown. The prevalence of hypertension in the population is shown by the dashed horizontal baseline line across the entire cohort per gene (4.98%). The statistical significance of the partition is indicated by asterisks for the dominant and recessive inheritance models. The sub-populations are specified by percentiles (e.g., “bottom 25%” refers to the 25% of the cohort with the lowest PWAS effect scores) where the partitioned is determined by selecting the cutoff with the lowest p-value by Fisher’s exact test. **(D)** Recessive (top) and dominant (bottom) effect of the PRS partition by the fold change of the two population as in (C). The contribution of males to the gene-based PRS is higher than that of the females for SH2B3.

The number of individuals for the best partition is shown for the dominant and recessive models. A quarter of the cases exhibited a substantial 1.24-fold difference in their risk for hypothyroidism. Interestingly, when we divided the analysis into males and females, we discovered that the contribution of genetics to phenotypes in males is stronger in almost all cases (**Figure 8D**), despite males having a lower overall prevalence than females (2.04% versus 7.45%, respectively). For example, in males, we identified 124 individuals that were associated with a >1.5-fold increased risk for the recessive inheritance model relative to the rest of the cohort (Supplementary **Table S7**) for DCLRE1B and 95 for DCF19. We confirmed that for all significant genes in PWAS, hundreds, and often thousands, of individuals can benefit from recording their genetics. The optimal partition of the relevant cohorts for selected genes by PRS-like analyses is reported in Supplementary **Table S7**. Each gene is analyzed separately for males, females, or both, and for a dominant or recessive inheritance model.

## Discussion

Hypothyroidism is a disease with a range of clinical manifestations and different causes ^46^. Primary hypothyroidism is measured by low levels of blood thyroid hormones. Hashimoto’s thyroiditis causes 90% of primary hypothyroidism cases, but other causes for the dysfunction of the thyroid gland (e.g., due to medication) are known. Less common are cases of secondary or tertiary hypothyroidism. Such conditions are caused by insufficient stimulation of the thyroid gland, often due to the failure of the pituitary gland. It may also be a result of dysfunction of the hypothalamus that fails to secrete enough thyrotropin-releasing hormone (TRH) to stimulate the pituitary gland. Other forms of hypothyroidism are associated with organ resistance to thyroid hormone ^47^. In addition, subclinical hypothyroidism, which mostly affects the aged population and especially females, is linked to insufficient iodine, which prevails worldwide ^48, 49^. A special case of subclinical hypothyroidism on the basis of pregnancy will not be further discussed ^50^.

An accepted assumption is that the serum measurements of TSH and free thyroxine (FT4) are genetically determined ^51^. We expect that the genetic basis for the increased level of TSH can potentially expose overlooked loci involved in hypothyroidism ^11, 52^. In this study, we investigate the genetic basis of hypothyroidism (ICD-10, E03) in the adult population of UKB. Under a routine GWAS methodology, variants are statistically tested for their effects under a case-control setting ^53^. GWAS is additive, whereas PWAS also detects non-additive effects such as recessive inheritance. In genetic terms, PWAS enables the detection of compound heterozygosity when both variants occur at different locations within the same gene. although only 12% of the PWAS genes exhibited clear recessive models. In general, recessive effects have been overlooked by the classical GWAS approach ^26, 54^. We conclude that both methods are complementary while revealing different aspects of hypothyroidism. Gene-based analysis was traditionally performed through the use of animal models, such as knockout mouse strains with thyroid receptor mutations ^55^.

We demonstrate that each gene-based association study method highlights a specific aspect of the disease etiology. Results from GWAS are available from independent cohorts and are often reported by summary statistics (e.g., as reported by OT, GWAS catalog). In classical GWAS for hypothyroidism, the most significant listed variants occur in the vicinity of genes that act in thyroid development and hormone secretion (e.g., PAX8, FOXE1, STAT4, TG; **Table 1, Figure 6B**). On the other hand, the complementary approach of TWAS seeks a signature of gene expression using tissue-based expression data. The significant variants from TWAS are located in regions that may alter the expression levels of transcripts, with cis or trans regulation modes ^56, 57^. We observed that TWAS results are quite different from those of the GWAS findings (**Figure 5C**), with both display only a modest signal of immunity ^58^. In the case of hypothyroidism, PWAS mostly identified the immunological signature of hypothyroidism ^58^. The predisposition variants across numerous autoimmune diseases ^59^ overlap with PWAS identified genes and variants of coding-GWAS. Testing the genetic variations associated with thyroid autoimmunity identified a strong interaction with the pathways driving the immune response toward PD-1 blockade in immunotherapy of lung cancer ^58, 60^. Coding variants in PTPN22, SH2B3, and numerous genes in the Class 1 MHC region can help predict the risk for autoimmune diseases and hypothyroidism ^11^. Autoimmune thyroiditis is associated with other immune-mediated diseases (T1D, Celiac, rheumatoid arthritis (RA), systemic lupus erythematosus (SLE), psoriasis and more). Hashimoto’s thyroiditis, which is the most common form of hypothyroidism, is characterized by infiltration of the T lymphocytes to the thyroid gland and autoantibodies against thyroid specific genes (e.g., thyroid peroxidase, thyroglobulin, and TSH receptor). The notion by which different genetic association methods expose complementary aspects of a disease will be tested for other complex polygenic diseases. We anticipate that the identification of different aspects of the disease etiology is feasible for diseases with congenital and acquired forms. It is likely that complex diseases for which the age of diagnosis (i.e., early onset and late onset) varies greatly may have an alternative mechanistic basis. Importantly, the association of lead variants with the appropriate causal gene is prone to inaccuracy due to linkage disequilibrium (LD) in highly polymorphic regions ^61^. Often, the reduction in false associations is carried out by fine-mapping, experimental evidence, and independent duplication (discussed in ^62^). To overcome inaccuracy in mapping, we have conducted a non-standard GWAS that relies on a set of variants solely confined to coding regions (the same set of variants as used for PWAS). Under this scheme, the dominant role of an immunological signature, mostly associated with chromosome 6, was revealed. Furthermore, both association methods (PWAS and coding-GWAS) include the same consideration of 172 covariates that account for population structure and numerous technical biases (see Methods). Using the coding-GWAS statistics for both sexes (at a threshold of absolute z-value >4), 64% of the genes were located in the MHC region (supplementary **Table S6)** which highlights the centrality of the MHC region to hypothyroidism in humans and mice ^63^. It allowed us to unequivocally identify genes that are highly enriched in immunity-related genes and specifically with the MHC region. Actually, even stronger effects, at a threshold of -log10p-value >10, listed for 115 variants (supplementary **Table S6**), among which as many as 93% were confined to a sub-region of the MHC that covers 2.4M bp in Chr6 (30.6M to 33.0M, **Figure 7C**).

While we were able to identify genes from different functional classes using multiple genetic association models, CH is mostly monogenic and reported by OMIM from exome sequencing with rare mutations ^64^. CH is frequently found in conjunction with other developmental phenotypes ^65^. For example, CH combined with neonatal diabetes mellitus is caused by mutations in the transcription factor (TF) GLIS3 gene ^66^. Other TFs include NR1D1, which coordinates circadian rhythm, and PAX8, which is exclusively expressed in the thyroid cell types. These genetic variants are often the cause of monogenic congenital hypothyroidism (CH). OMIM reports on 34 such CH genes (Supplementary **Table S8**) that includes key TFs such as FOXE1 and STAT4. These genes act during embryogenesis to establish the pituitary, hypothalamus, and thyroid axis (**Table 1**). Combining results from multiple classical GWAS identified TFs that have strong links to thyroid function (e.g., NKX2-1; NK2 homeobox 1). Thyroid signaling genes are also strongly represented in TWAS. For example, PDE8B is expressed primarily in the thyroid to execute the TSH effects. Interestingly, these thyroid-related TFs prevail in the GWAS list and in the gene overlap with TWAS findings (**Figure 6C**). However, none of the top GWAS high-scoring OT genes were identified by PWAS (**Figure 6B**). We attribute this discrepancy in gene finding to the relatively small effect size of most common variants in coding genes. For the rest of the genome, GWAS allowed significant non-coding variants to be identified. Moreover, GWAS potentially identifies functional elements that are ignored by PWAS or coding-GWAS. For example, a cluster of variants on chromosome 9 including rs10759927, rs7850258 and rs7030280, was significantly identified by classical GWAS for hypothyroidism (with p-value ranging between 1e-82 to 2e-100). These variants occur are located within the introns of PTCSC2, a non-coding transcript (ncRNA) that is expressed exclusively in the thyroid.

It is critical to validate the results of any association studies by independent cohorts. To this end, we used the large-scale genotyping of FinnGen (Fz7). A limitation in comparing numerous genetic association methods is due to the slightly different gene sets covered by the different resources (e.g., OT, TWAS, PWAS, coding-GWAS). For example, the complete human proteome includes 20,389 genes; among them, about 2,000 genes are not included in the analyzed gene list for PWAS (see Supplementary **Table S1**). We noted that several of the hypothyroidism associated genes reported by FinnGen are not covered by either coding-GWAS or PWAS and thus could not be validated. These genes are DIO1, HSP89, NINJ2, PTPN22, SYN2, TMEM71, and TSHR. Notably, PTPN22, SYN2 and TSHR genes were also reported among the top scoring genes in OT (**Figure 6B)**. Genes that failed in mapping (see Methods) will be addressed in future versions of PWAS.

The gene-based analysis performed in this study raised the question of whether the effect size of genetic variants is distinguished by sex. A ratio of > 3:1 for females and males was reported in UKB and in FinnGen Fr7. However, the overall occurrence in the Finnish population is higher, with 18% in females (37,942 cases) and in 5.8% males (9,616 cases). The average diagnosis age also differs with 50.0 and 58.3 years in females and males, respectively. Despite differences in sex prevalence ^67^, most human traits and diseases, excluding immune-related diseases, do not support a mechanistic difference between the sexes ^68^. With the continuous increase in the statistical power due to cohort sizes, more cases of sex-dependent genetics are revealed. For example, significant sex-dependent effects were enriched for neuronal development and the immune-related genes in the cases of schizophrenia, major depression disease ^69^, autism ^70^ and Alzheimer’s disease ^71^. It was proposed that the apparent sex difference might be due to participation bias (e.g. indirect effect of sex-differential mortality) ^72^. In this study, we showed that the sex stratification for hypothyroidism provided no support for genuine genetic effect differences between the sexes, despite a ratio of 3:1 in the disease occurrence in females to males.

## Data Availability

All data in the present study are based on the UKB for application application ID 26664 (Linial lab). The code, parser and filtration scheme used for the analyses are available in GitHub of the Linial lab.

https://github.com/nadavbra/pwas

## Acknowledgements

We thank Naomi van Wijk for reading the manuscript and for her insightful comments. We thank Dr. Nadav Brandes for developing the UKB parser. We thank Dr. Guy Kelman for his help in the implementation of PWAS. We thank the Linial’s lab for useful discussions. We appreciate the constant support of the system team of the School of Computer Science at the Hebrew University.

## Data and code availability

The UK-Biobank (UKB) application ID 26664 (Linial lab). Ethical committee approval, The Hebrew University #13082019 from the University Committee for the Use of Human Subjects in research (dated 07/2002). PWAS is available through command-line interface as part of an open-source project (MIT license) at Brandes N. pwas. Github. 2020. https://github.com/nadavbra/pwas. Accessed 11 Apr 2020.

## Supplementary data

Figures S1-S4; Tables S1-S8.

Table **S1**. A list of protein coding genes that are not included in PWAS analysis (total 2119). **Table S2**. Summary statistics for hypothyroidism comparative study by classical multiple GWAS. **Table S3**. Results of PWAS with 95 significant genes by their effect size and heritability modes genes (q-value <0.1). **Table S4**. A sorted list of 715 genes identified by genetic associations from OT. **Table S5**. Results from 100 PWAS runs for males and females following resampling. **Table S6**. Table of significant sex-dependent coding-GWAS. **Table S7**. Summary of gene-based PRS according to heritability models and sex. **Table S8**. A list of genetic related genes for CH according to OMIM.

## Supplementary Figures

**Figure S1.**
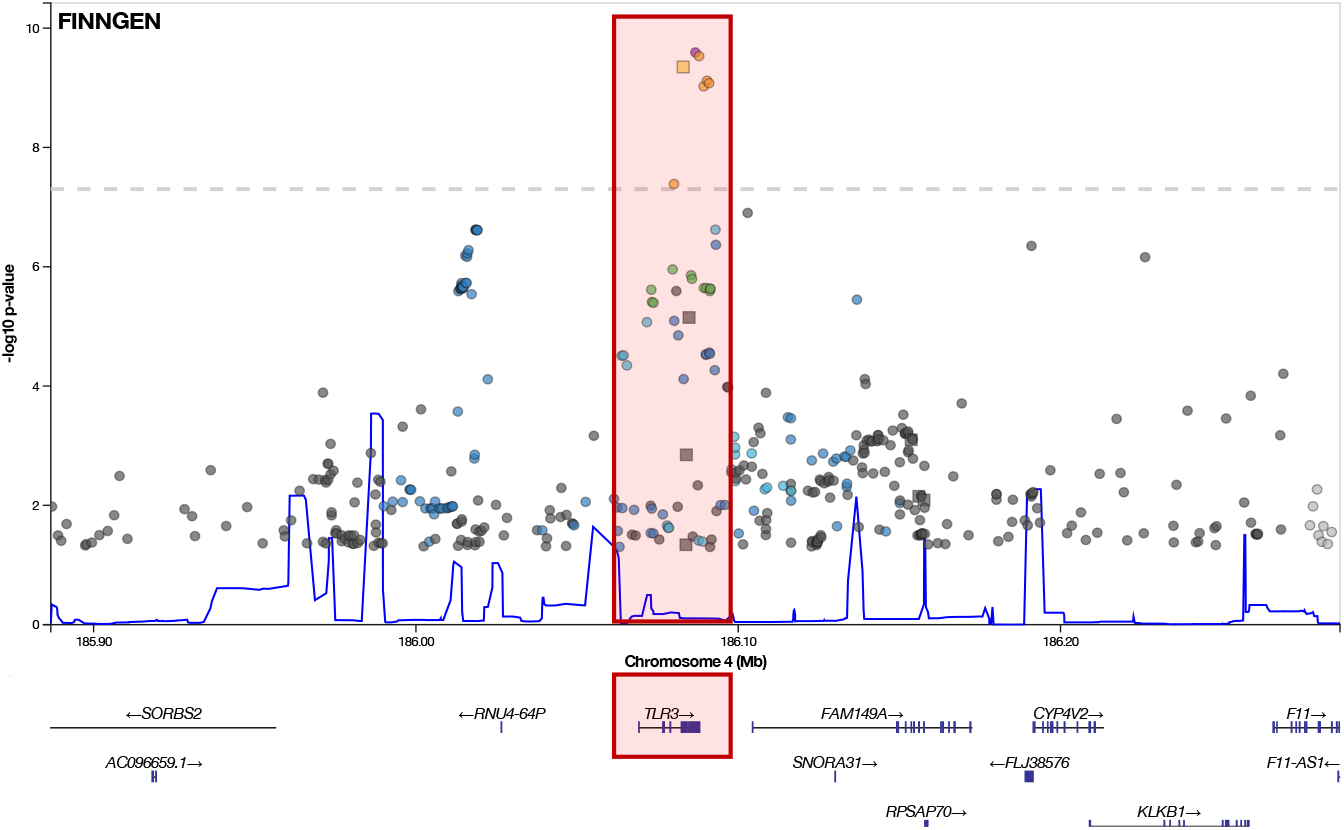
Fine mapping of loci from Chromosome 4 from 185.9 to 186.3 Mbp. TLR3 is the candidate causal gene indicated by the red frame. Data is extracted from FinnGen Fr7.

**Figure S2.**
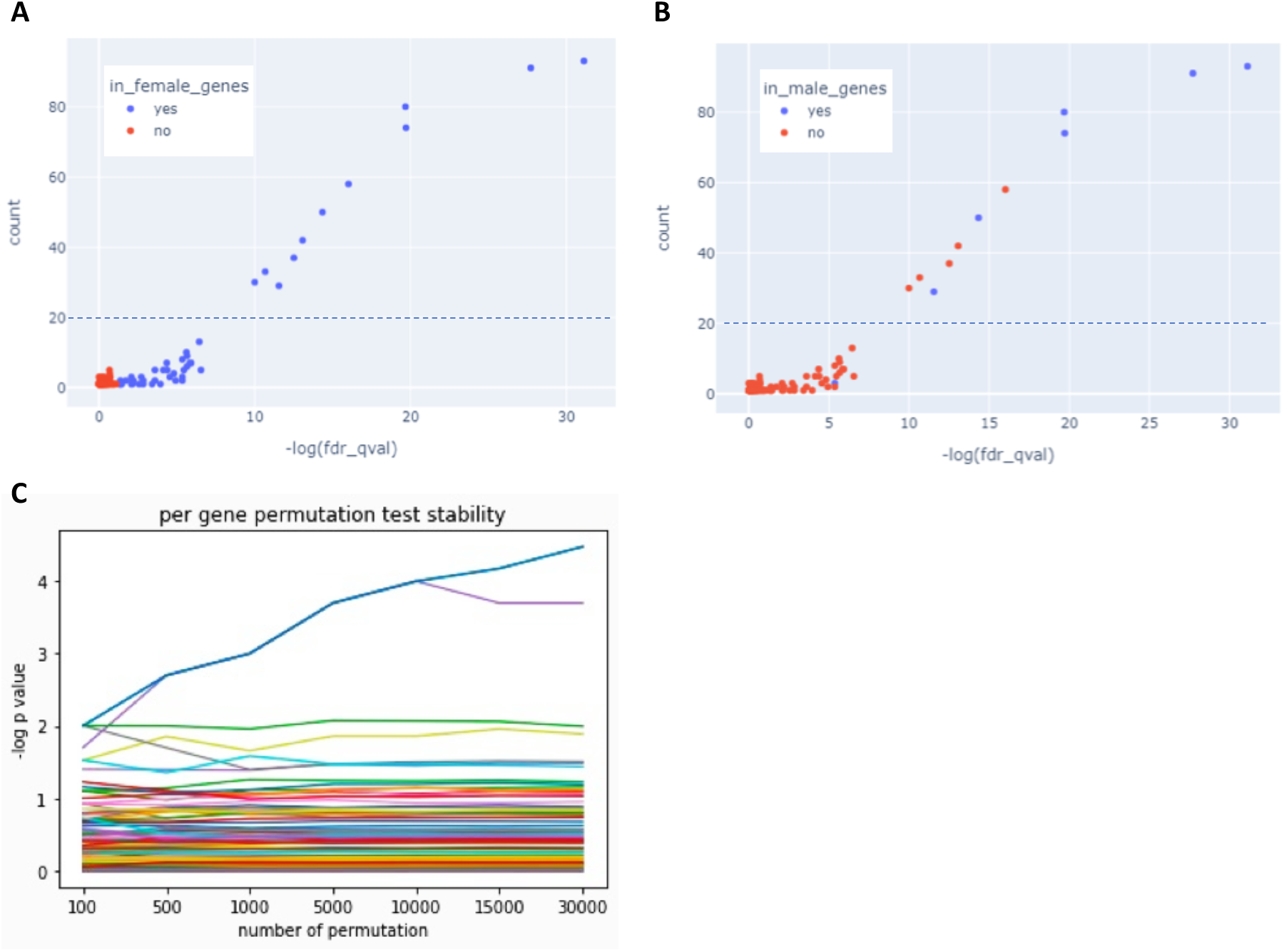
Testing sex-specific genetic-based signal by resampling and reduce the number of females by the size of the male group. The number of occurrences for a significant gene is shown across 100 independent PWAS runs. The number of occurrences for a significant gene is shown across 100 independent PWAS runs. **(A)** Colored by the PWAS results for females. **(B)** Colored by the PWAS results for males. The blue dots indicate genes that were initially included as significant in PWAS. The red dots are identified genes that were not significant in the full cohort PWAS analysis. For each sex, a handful genes were repeatedly identified in both sexes that are genuine to hypothyroidism. The other genes (20% occurrences, dashed line) are likely to be robust to the random noise. **(C)** Results along 30,000 permutation tests where the label of the sex is permuted. The test shows that 2 genes (and another 2 at lower confidence) reached statistical significance (i.e. differs substantially between males and females).

**Figure S3.**
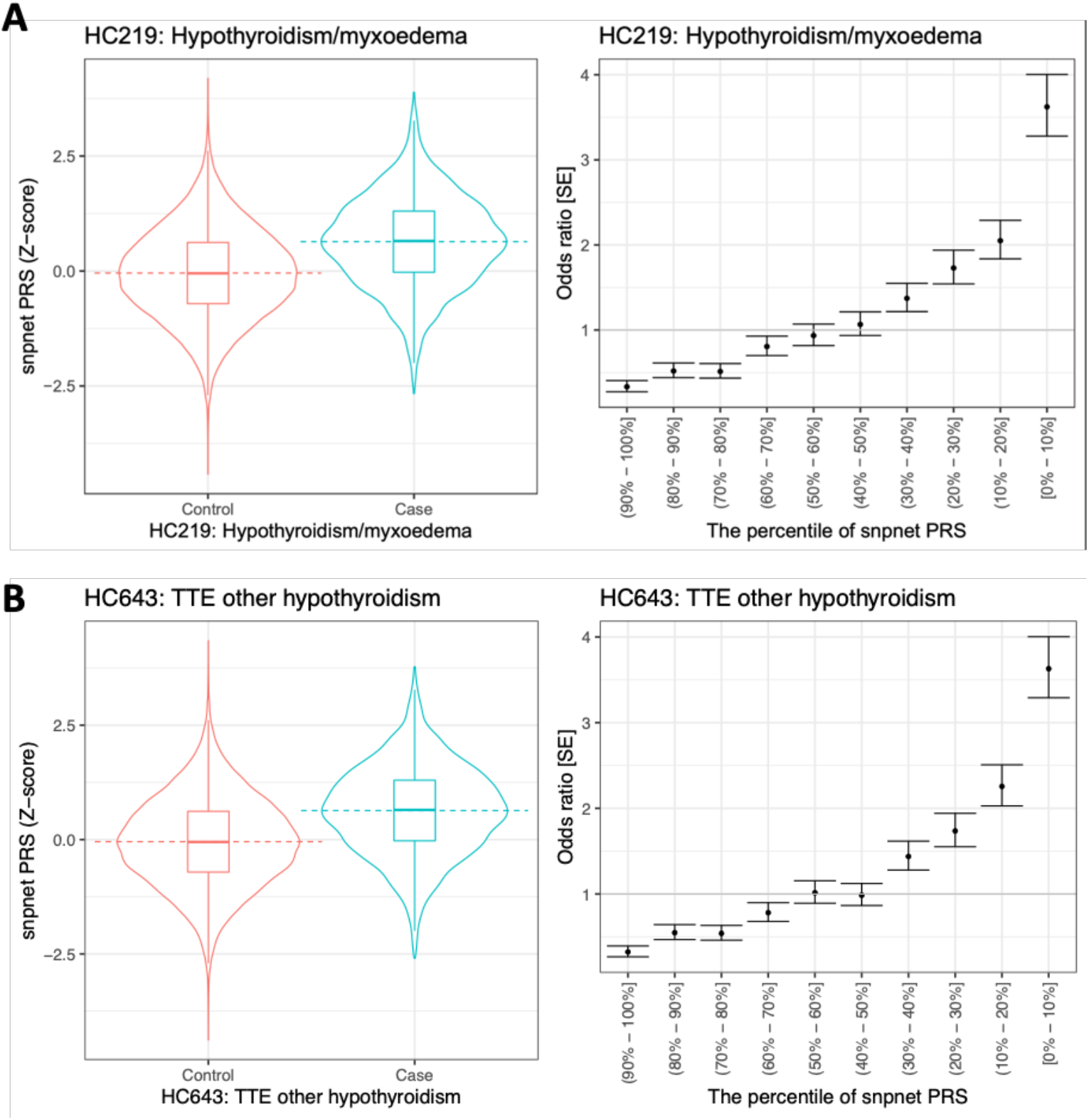
PRS for hypothyroidism/myxedema and TTE (time to event) hypothyroidism. Data was extracted from the Global Biobank engine ^45^. The analysis of the snpnet PRS model was developed using training set (i.e., model development set) of white British ancestry from UKB. The predictive performance was evaluated on a hold-out test set of individuals from the same origin.

**Figure S4.**
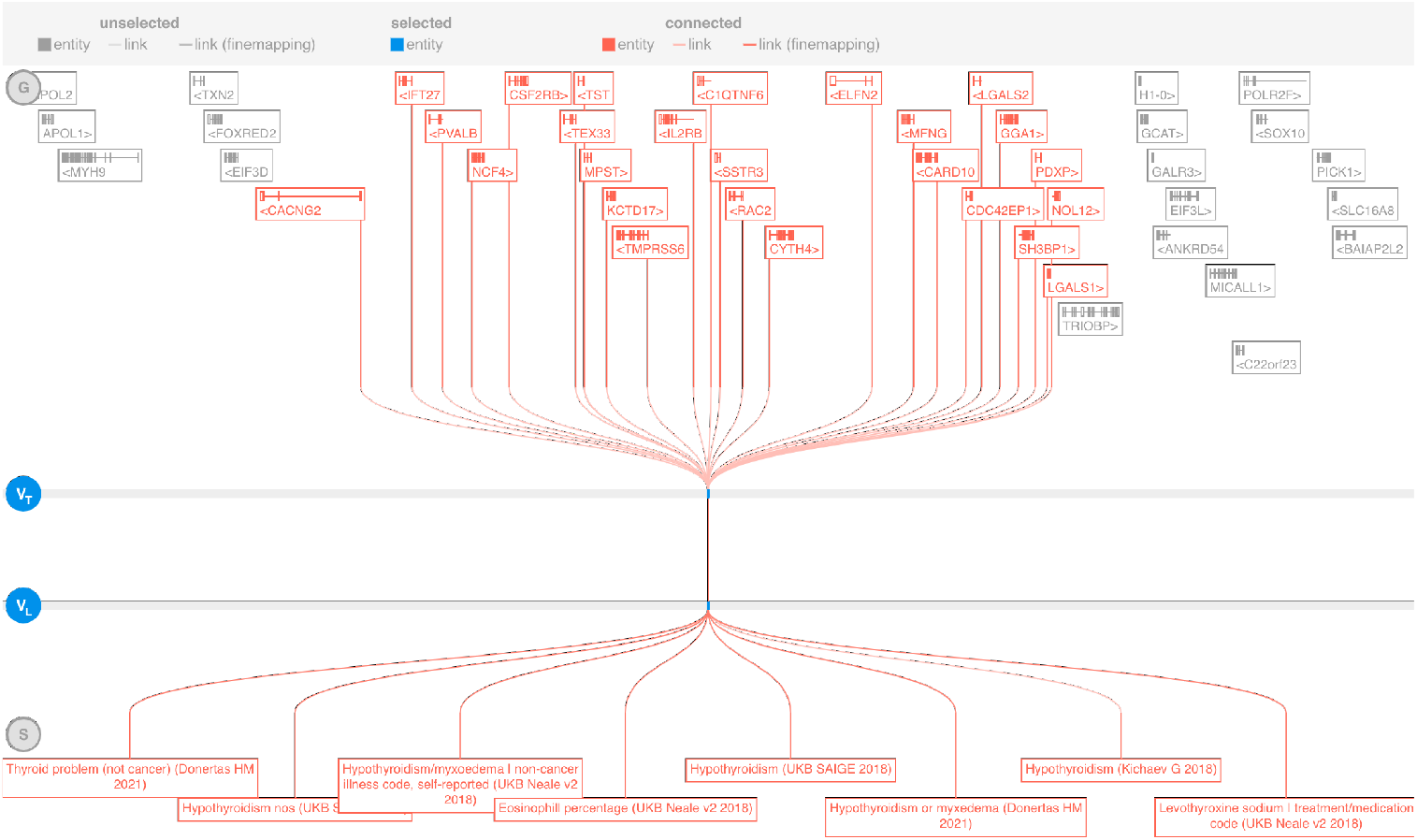
Loci around the gene C1QTNF6 (C1q and TNF related 6) that is located in the vicinity of >20 genes that could not be determined explicitly as causal. Note that genes within the loci are associated with hypothyroidism and several thyroid-specific phenotypes.

## Notes

### Competing Interest Statement

The authors have declared no competing interest.

### Funding Statement

ISF Grant 2753/20 to M.L. on Sex dependent genetics.

### Author Declarations

Ethics committee of the Hebrew University of Jerusalem gave ethical approval for this work.

